# A phase I/II clinical trial of intradermal, controllable self-replicating RNA vaccine EXG-5003 against SARS-CoV-2

**DOI:** 10.1101/2023.10.07.23296699

**Authors:** Takenao Koseki, Mayumi Teramachi, Minako Koga, Minoru S.H. Ko, Tomokazu Amano, Hong Yu, Misa Amano, Erica Leyder, Maria Badiola, Priyanka Ray, Jiyoung Kim, Akihiro C. Ko, Achouak Achour, Nan-ping Weng, Takumi Imai, Hisako Yoshida, Satsuki Taniuchi, Ayumi Shintani, Hidetsugu Fujigaki, Masashi Kondo, Yohei Doi

## Abstract

mRNA vaccines against severe acute respiratory syndrome coronavirus 2 (SARS-CoV-2) have played a key role in reducing morbidity and mortality from coronavirus disease 2019 (COVID-19). We conducted a double-blind, placebo-controlled phase I/II trial to evaluate the safety, tolerability, and immunogenicity of EXG-5003, a two-dose, controllable self-replicating RNA vaccine against SARS-CoV-2. EXG-5003 encodes the receptor binding domain (RBD) of SARS-CoV-2 and was administered intradermally without lipid nanoparticles (LNP). The participants were followed for 12 months. Forty healthy participants were enrolled in Cohort 1 (5 µg per dose, n = 16; placebo, n = 4) and Cohort 2 (25 µg per dose, n = 16; placebo, n = 4). No safety concerns were observed with EXG-5003 administration. SARS-CoV-2 RBD antibody titers and neutralizing antibody titers were not elevated in either cohort. Elicitation of antigen-specific cellular immunity was observed in the EXG-5003 recipients in Cohort 2. At the 12-month follow-up, participants who had received an approved mRNA vaccine (BNT162b2 or mRNA-1273) >1 month after receiving the second dose of EXG-5003 showed higher cellular responses compared to equivalently vaccinated participants in the placebo group. The findings suggest a priming effect by EXG-5003 on the long-term cellular immunity of approved SARS-CoV-2 mRNA vaccines.

## Introduction

Since its onset in late 2019, the COVID-19 pandemic has resulted in over 7 million deaths ^1^ and has left many more people with negative long-term health consequences worldwide. Against this backdrop, the rapid development of multiple COVID-19 vaccines has been a significant achievement in the field of biomedical research, with billions of vaccine doses distributed and administered worldwide ^2^. In particular, synthetic mRNA vaccines made a noteworthy debut following extensive fundamental and preclinical investigations that preceded the pandemic ^3,4^. With their rapid development, cell-free manufacturing, and excellent clinical efficacy, mRNA vaccines surpassed conventional live-attenuated, inactivated, and protein-based subunit vaccines in the race for early vaccine approval. Nonetheless, mass administration of mRNA vaccines has also revealed challenges, including adverse effects and the need for multiple booster doses. Another consideration in vaccine development is the role of vaccine-induced cellular immunity in protection against severe disease, hospitalization, and death. As SARS-CoV-2 variants that evade neutralizing antibodies continue to emerge, the significance of cellular immunity, especially CD8+ T cells, in safeguarding against severe disease in the long run is increasingly appreciated ^5–7^. In contrast to the limited cross-reactivity of neutralizing antibodies induced by vaccines against variants, T cell responses elicited by vaccines demonstrate excellent cross-reactivity. However, the approved mRNA vaccines induce weak cellular immunity, especially in CD8+ T cells ^8–10^, that wanes rapidly along with humoral immunity ^11,12^, though some groups report longer-lasting T cell immunity ^13–15^.

The weak induction and rapid waning of cellular immunity of the currently approved mRNA vaccines could be overcome by another type of mRNA vaccines using self-replicating RNA (srRNA), also called self-amplifying RNA (saRNA or SAM-RNA) ^16^, as it is known to be a strong inducer of cellular immunity in animal studies ^17–23^. However, clinical trials of the SARS-CoV-2 vaccine based on the srRNA/saRNA platform thus far have shown lower than expected immunogenicity in humans ^24–26^.

The EXG-5003 vaccine is based on srRNA/saRNA technology, but it is engineered to be controllable by temperature: it functions at around 33°C – skin temperature, but does not function at 37°C – core body temperature ^27^. Thus, this novel RNA vaccine platform is called controllable self-replicating RNA (c-srRNA) ^27^. Unlike mRNA and srRNA/saRNA vaccines that are administered to skeletal muscles, the c-srRNA vaccine platform is optimized for intradermal delivery, which may enhance cellular immunity as it is well known that intradermal vaccination can induce strong cellular immunity ^28,29^. Also, unlike mRNA and srRNA/saRNA vaccines that are complexed with LNP, the c-srRNA vaccine platform is not complexed with LNP and is delivered as naked RNA, which may reduce local reactogenicity in the skin, which has often been an issue for previous intradermal vaccines. Preclinical studies have shown that intradermal vaccination with EXG-5003 alone induced robust cellular immunity but did not induce humoral immunity ^27^. However, EXG-5003 showed priming effects, as demonstrated by a rapid and strong induction of humoral immunity after subsequent exposure to the same or variant RBD antigen compared with control ^27^.

Here, we report the results of a phase 1/2 study to evaluate the safety, tolerability, and immunogenicity of EXG-5003 in healthy adults.

## Results

### Participants

Of 46 participants who gave consent, 40 participants met the inclusion criteria. They were enrolled in Cohort 1 and Cohort 2 of this study, with 16 participants assigned to receive EXG-5003 and 4 participants to receive placebo in each cohort, respectively (Figure 1). In Cohort 1, one participant in the EXG-5003 5 µg group did not receive the second dose due to an unrelated injury requiring hospitalization. All 40 participants completed follow-up to Day 57. One and 3 participants in Cohort 1 (EXG-5003 5 µg group) and Cohort 2 (EXG-5003 25 µg group) did not return at the 1-year final follow-up, respectively.

**Figure 1.**
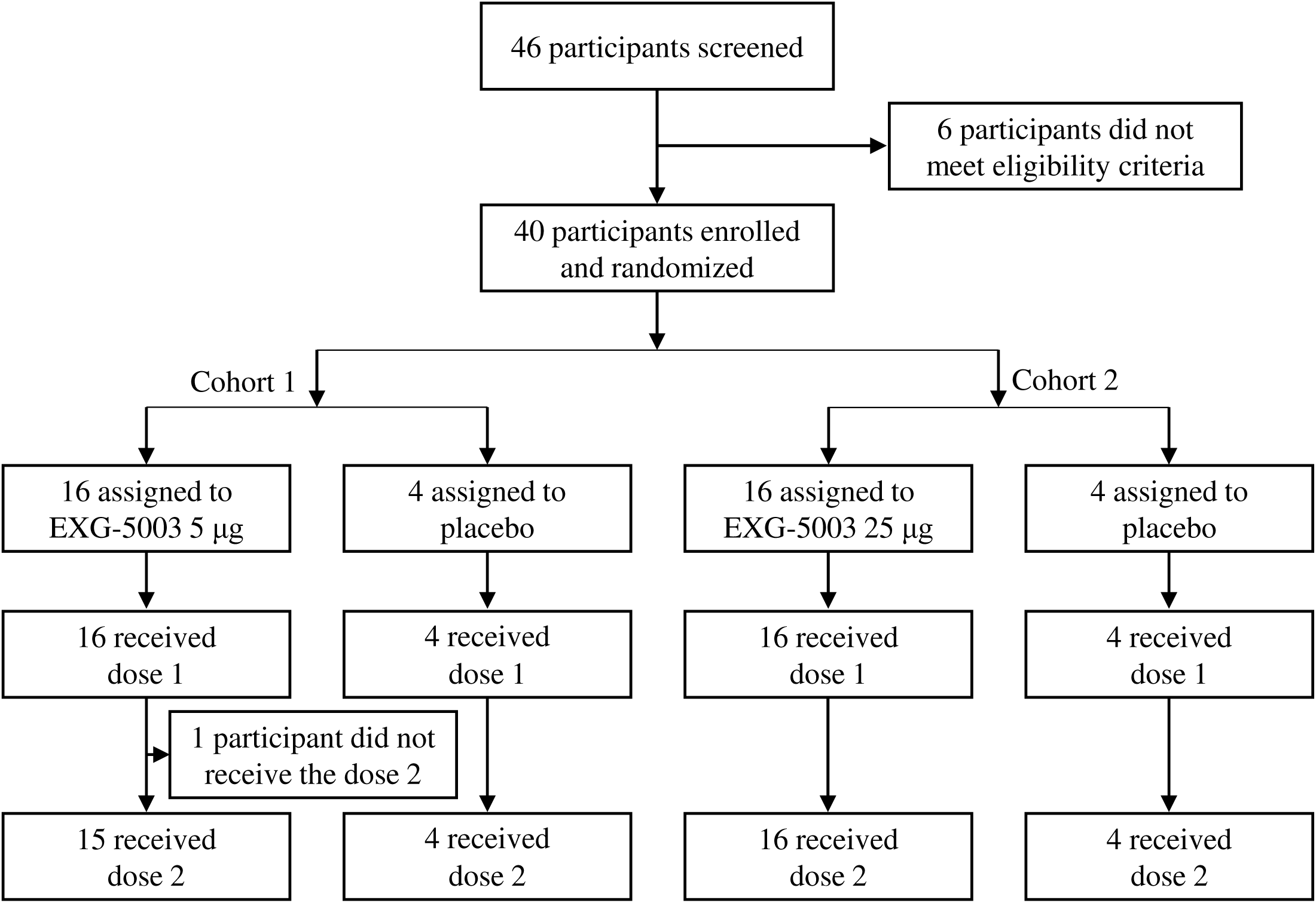
A flow chart showing enrollment and randomization of participants. Cohort 1 (5 µg group) was dosed first. After the second dose, the safety was monitored for two weeks. Then, Cohort 2 (25 µg group) was dosed.

The participant backgrounds for the full analysis set (FAS) in this study are shown in Table 1. For Cohort 1, males accounted for 10/16 (62.5%) in the EXG-5003 5 µg group and 3/4 (75%) in the placebo group. The median age was 39.5 years in the EXG-5003 5 µg group and 48 years in the placebo group. For Cohort 2, males accounted for 7/16 (43.8%) in the EXG-5003 25 µg group and 4/4 (100%). The median age was 43 years in the EXG-5003 25 µg group and 44 years in the placebo group.

**Table 1.**
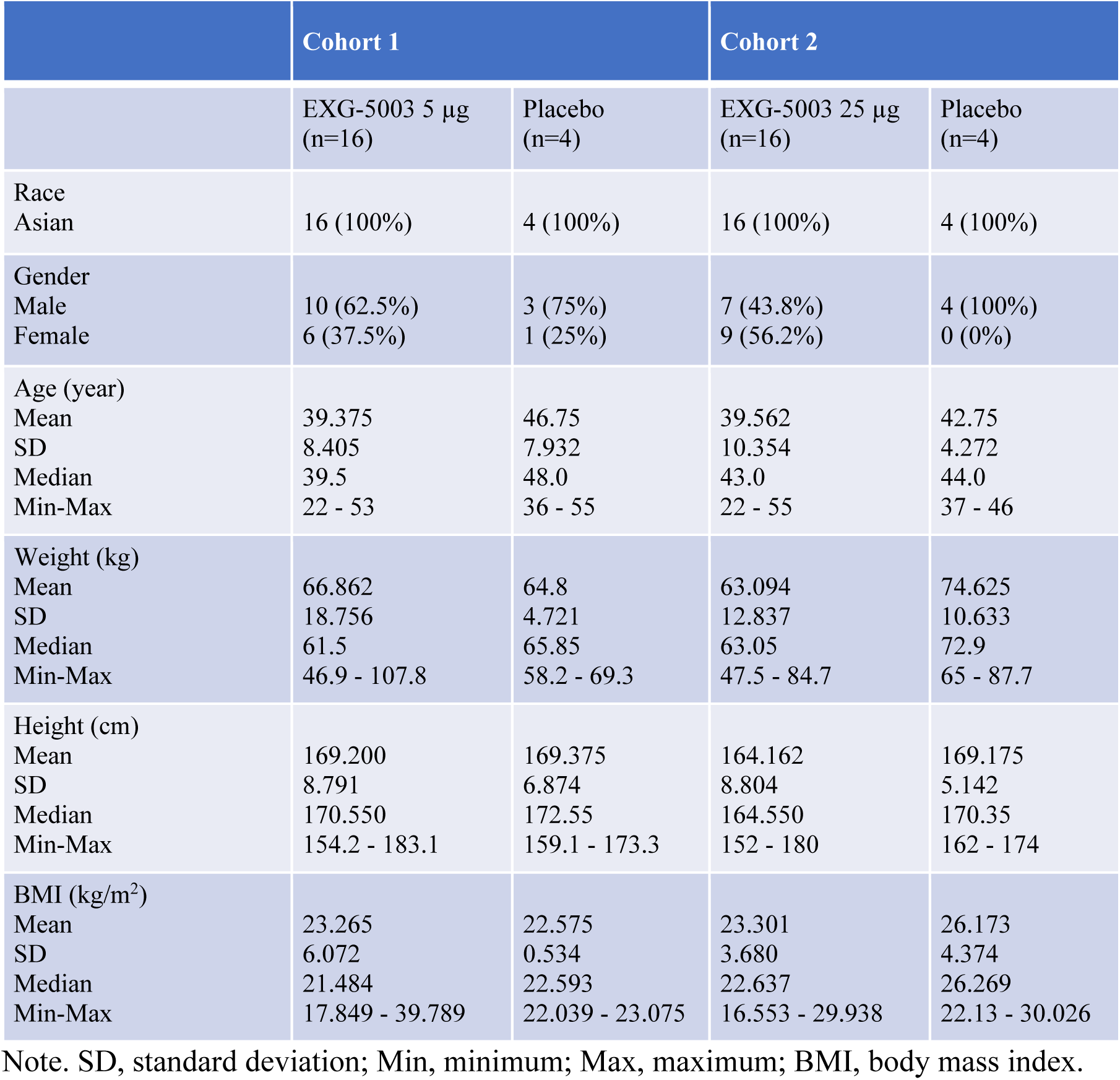
Participants demographics.

### Safety

Local adverse events or reactogenicity associated with the administration of EXG-5003 are summarized in Figure 2A. Only Grade 1 events were reported from a small number of participants. A photo shows an example of observed redness (Figure 2A). Systemic adverse events are summarized in Figure 2B. Only minor events, mostly Grade 1 and some Grade 2, were reported. It is noteworthy that the reactogenicity and systemic adverse events associated with EXG-5003 were much lower than the approved mRNA vaccines ^3,4^ and other srRNA/saRNA vaccines ^26,30^. This low reactogenicity of the intradermal EXG-5003 vaccine was expected because the vaccine contains only RNA and did not contain skin irritants such as lipids or oils.

**Figure 2.**
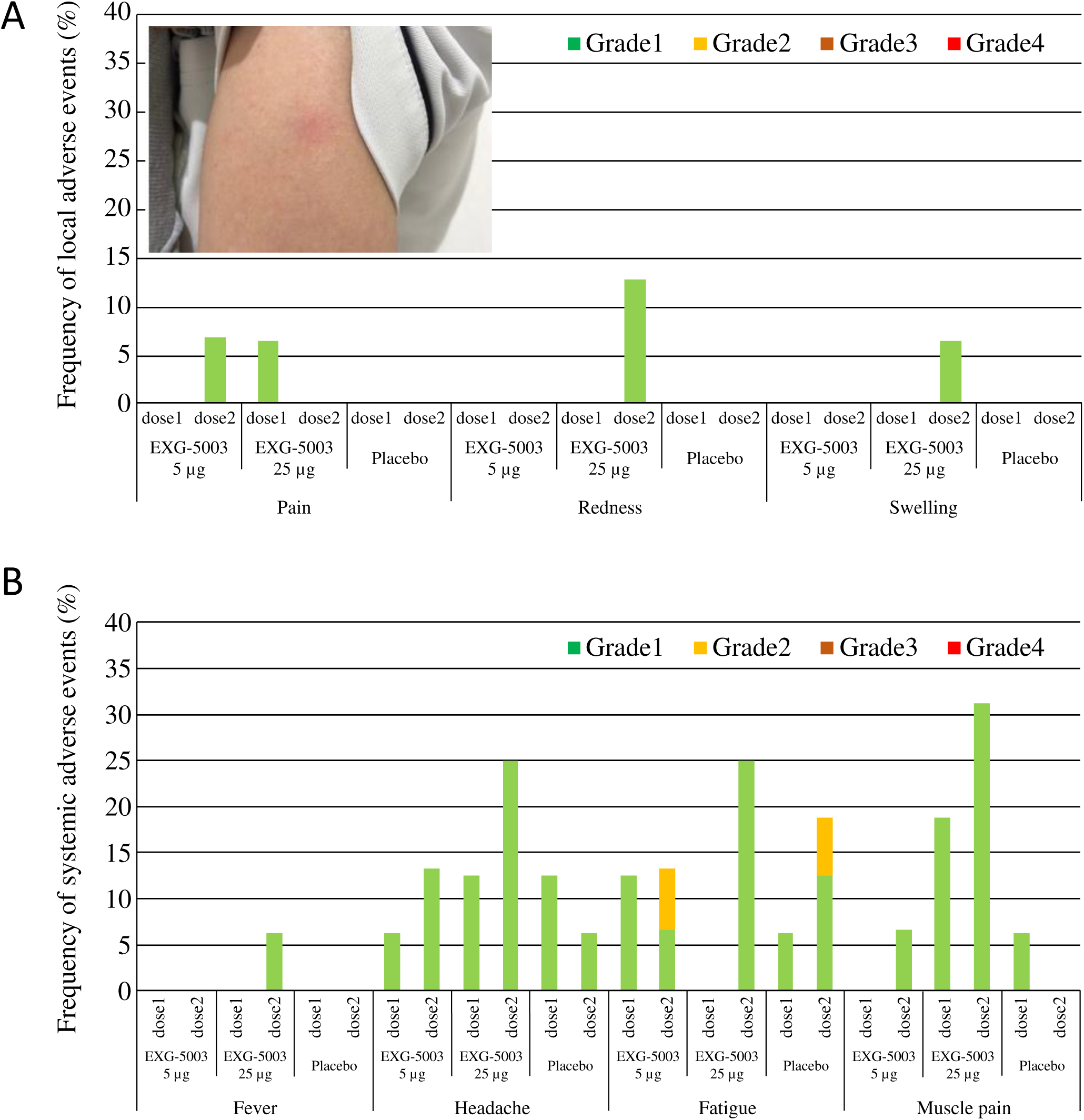
Safety of EXG-5003. (A) Frequency of local adverse events within 7 days of vaccination and a photo example of redness at the intradermal injection site. (B) Frequency of systemic adverse events within 7 days of vaccination.

### Short-term humoral immunity elicited by EXG-5003

The results of the FAS for anti-SARS-CoV-2 RBD antibody titers are shown in Figure 3, which plots the geometric means of titer for each participant by dose group and by time of evaluation over time (Table S1). Placebo groups consisting of 4 participants each from Cohort 1 and Cohort 2 were plotted together. As external controls, serum from four convalescent individuals who were hospitalized for COVID-19 and one serum sample from the WHO COVID-19 reference were analyzed in parallel (Table S2). Because the participants were allowed to receive licensed COVID-19 vaccines after Day 57, humoral immunity elicited by EXG-5003 alone could be assessed until Day 57. As shown in Figure 3, some individuals in the placebo group showed antibody titers higher than those who were vaccinated at some time points. They were negative for COVID-19 based on ELISA results against the nucleoprotein of SARS-CoV-2 and did not receive licensed COVID-19 vaccines during this period; thus, the reason for these high titers was not clear. For EXG-5003 vaccinated groups, low or undetectable levels of an anti-SARS-CoV-2 RBD antibody were observed (Figure 3A). Similarly, the results of the FAS for neutralizing antibody (ntAb), by both cell-free SARS-CoV-2 neutralization assay (Figure 3B, Table S3) and pseudotyped lentivirus assay (Figure 3C, Table S4), showed low or no increase of titers compared to the placebo group. Data points shown in red indicate individuals who received at least one dose of the approved mRNA vaccines (BNT162b2 or mRNA-1273) at the time of serum collection. These data will be discussed later.

**Figure 3.**
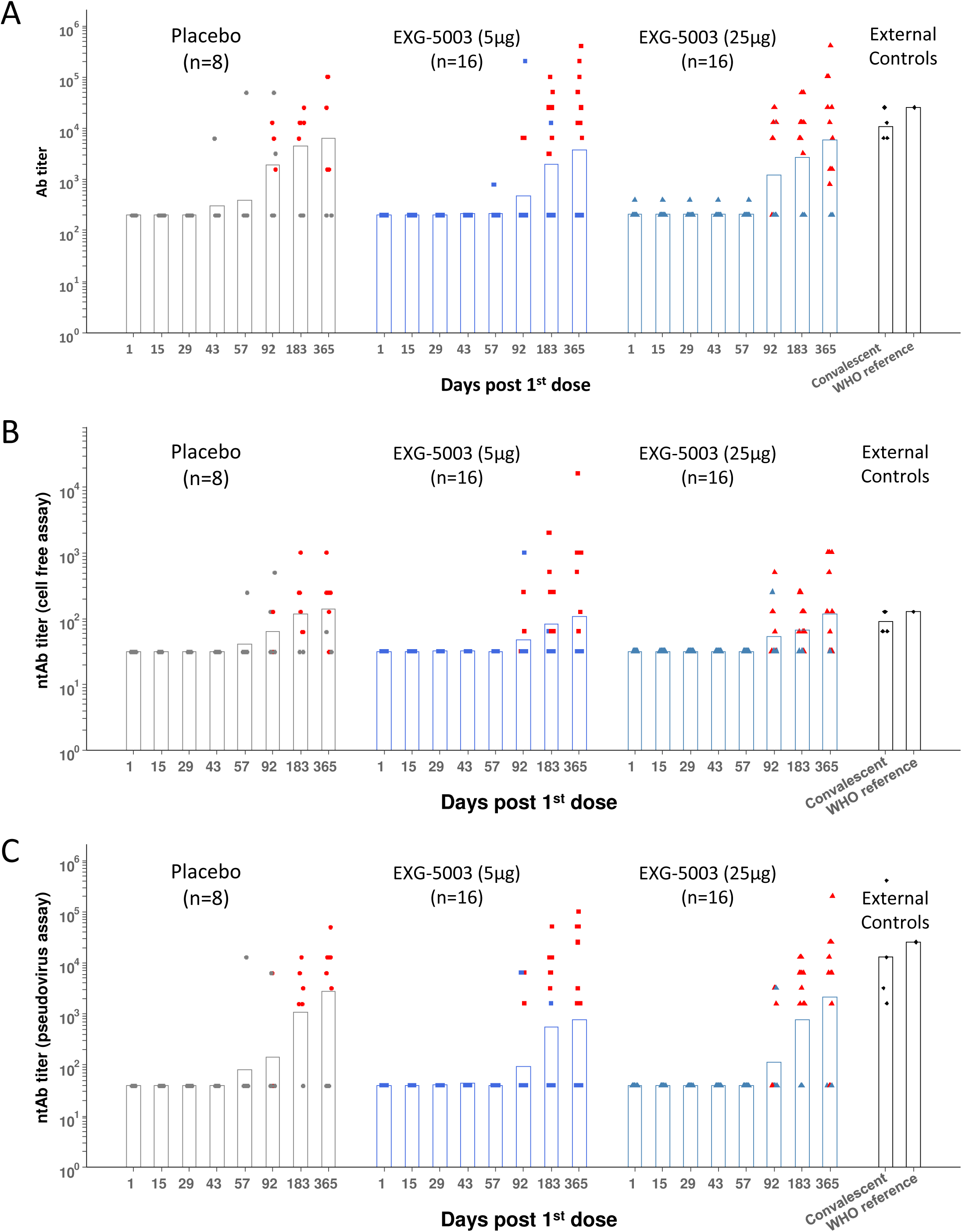
Humoral immune responses against the RBD of SARS-CoV-2. Geometric means of antibody titers. (A) Geometric means of anti-RBD antibody titer. (B) Geometric means of neutralization antibody titer by cell-free SARS-CoV-2 neutralization assay. (C) Geometric means of neutralization antibody titer by the SARS-CoV-2 spike pseudo-typed lentivirus assay. Red color indicates participants who received at least one dose of approved mRNA vaccine (BNT162b2 or mRNA-1273). FAS, the full analysis data set.

### Short-term cellular immunity elicited by EXG-5003

The results of the FluoroSpot assay for IFN-γ and IL-4 are shown in Figure 4. Cellular immunity was tested by stimulating PBMCs with a 15-mer peptide pool and a 9-mer peptide pool, which cover the RBD antigen. Short-term cellular immunity (up to Day 57) did not seem to be induced by EXG-5003 compared to the placebo and external controls, including convalescent individuals and individuals vaccinated with the BNT162b2 mRNA vaccine (Figure 4). However, some individuals in the placebo group and one individual in the external controls before BNT162b2 vaccination (BNT16b2 [pre]) showed cellular immunity comparable to the convalescent individuals. Also, two of three individuals who received one dose of BNT162b2 vaccine (BNT162b2 [1 dose]) and one of three individuals who received two doses of BNT162b2 vaccine (BNT162b2 [2 doses]) showed rather low cellular immunity, which is comparable to the EXG-5003 vaccinated group. Measurements for Cohort 1 (16 EXG-5003 5 μg and 4 placebo participants) and Cohort 2 (16 EXG-5003 25 μg and 4 placebo participants) were conducted in different batches on separate periods. We reasoned that the assays performed at the same time would demonstrate more consistent results. Therefore, we looked at Cohort 2 separately (Figure 5). This showed a clearer pattern of gradual increase of cellular immunity in the EXG-5003 group compared to the placebo group (Figure 5). Notably, some individuals who received EXG-5003 25 µg even after a single dose showed higher cellular immunity than two of three individuals who received a single dose of BNT162b2 vaccine (BNT162b2 [1 dose]) and one of three individuals who received two doses of BNT162b2 vaccine (BNT162b2 [2 doses]).

**Figure 4.**
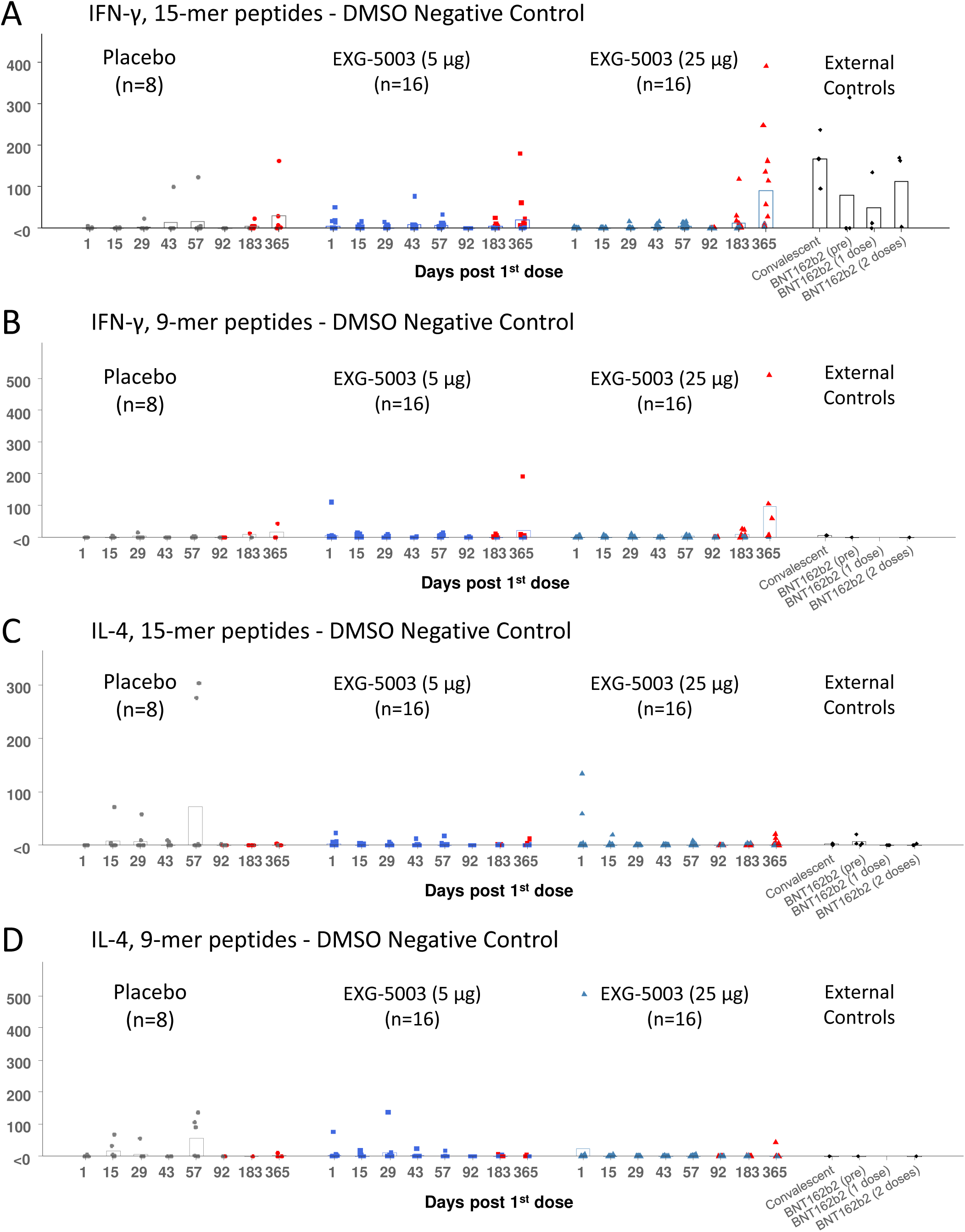
Cellular immune responses against the RBD of SARS-CoV-2. (A) IFN-γ spot-forming units (SFU) per 10^6 PBMCs stimulated by 15-mer RBD peptide library. (B) IFN-γ SFU per 10^6 PBMCs stimulated by 9-mer RBD peptide library. (C) IL-4 SFU per 10^6 PBMCs stimulated by 15-mer RBD peptide library. (D) IL-4 SFU per 10^6 PBMCs stimulated by 9-mer RBD peptide library. Red color indicates data points after receiving at least one dose of approved mRNA vaccine (BNT162b2 or mRNA-1273). External controls (for 15-mer RBD peptide library): Convalescent (n=3, blood was collected 20, 21, 22 days after the onset of symptoms, respectively), BNT162b2 vaccine (pre) (n=4, before receiving 1st dose of BNT162b2), BNT162b2 (1 dose) (n=3, two weeks after receiving the 1st dose of BNT162b2), BNT162b2 vaccine (2 doses) (n=3, two weeks after receiving the 2nd dose of BNT162b2). External controls (for 9-mer RBD peptide library): Convalescent (n=1, blood was collected 20 days after the onset of symptoms, respectively), BNT162b2 vaccine (pre) (n=1, before receiving 1st dose of BNT162b2), BNT162b2 vaccine (2 doses) (n=1, two weeks after receiving the 2nd dose of BNT162b2).

**Figure 5.**
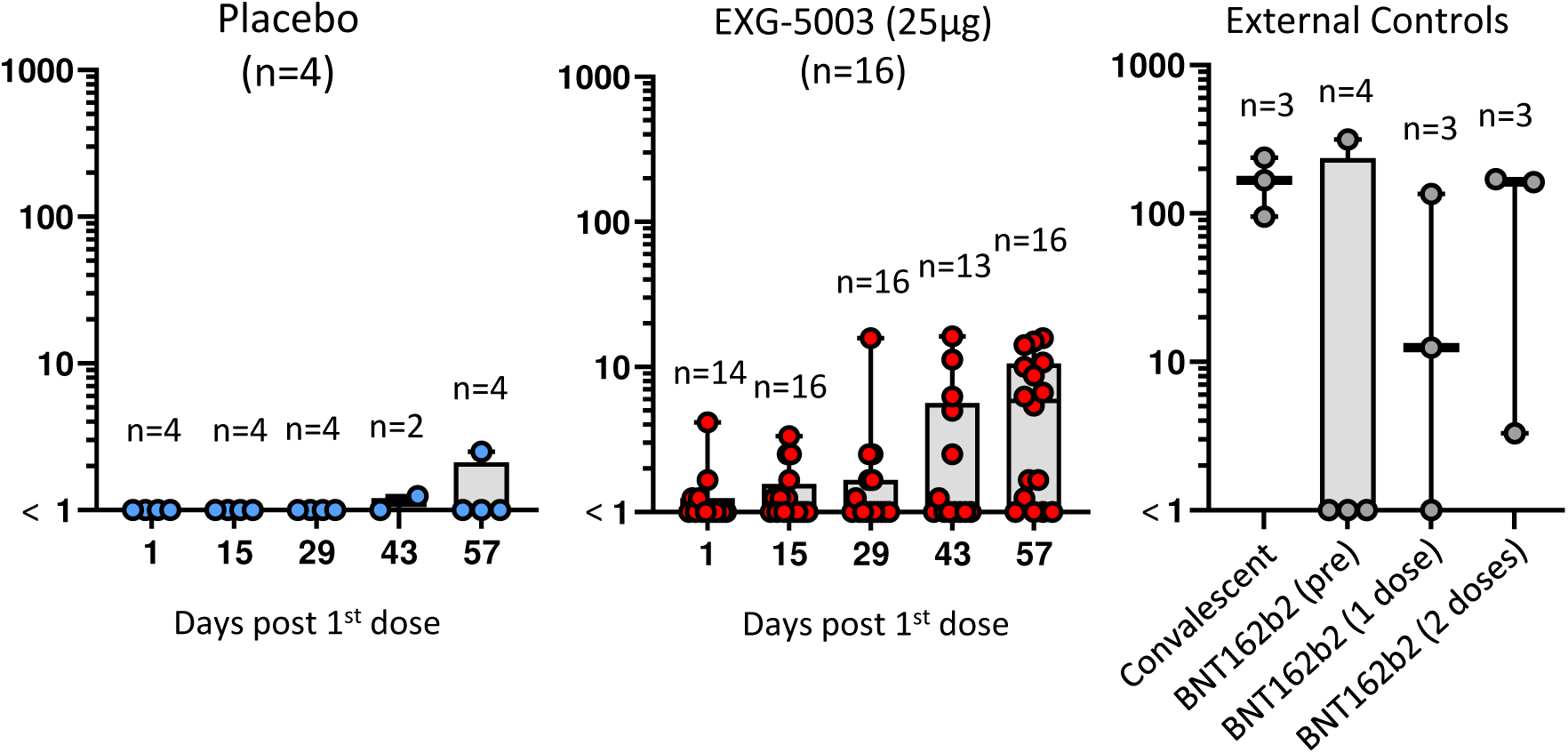
Cohort 2 cellular immune response data replotted from Figure 4 by log-scale. IFN-γ spot-forming units (SFU) per 10^6 PBMCs stimulated by 15-mer peptide library. Placebo (n=4) in blue dots (left). EXG-5003 (25 µg, n=16) in red dots (middle). External controls: Convalescent (n=3, blood was collected 20, 21, 22 days after the onset of symptoms, respectively), BNT162b2 vaccine (pre) (n=4, before receiving 1st dose of BNT162b2), BNT162b2 vaccine (dose 1) (n=3, two weeks after receiving the 1st dose of BNT162b2), BNT162b2 vaccine (2 doses) (n=3, two weeks after receiving the 2nd dose of BNT162b2). External controls (for 9mer peptide library): Convalescent (n=1, blood was collected 20 days after the onset of symptoms, respectively), BNT162b2 vaccine (pre) (n=1, before receiving 1^st^ dose of BNT162b2), BNT162b2 vaccine (2 doses) (n=1, two weeks after receiving the 2nd dose of BNT162b2).

### Long-term humoral immunity elicited by EXG-5003 following administration of licensed vaccines

During this trial, approved SARS-CoV-2 vaccines became available in Japan. Therefore, the participants were unblinded from randomization at Day 57 and allowed to receive the approved SARS-CoV-2 vaccines (BNT162b2, mRNA-1273, ChAdOx1, NVX-CoV2373) during the rest of the follow-up period. Some participants started to receive the approved vaccines. At the time of the final 1-year follow-up, some participants took two or three doses of the approved vaccines (BNT162b2 or mRNA-1273), whereas some participants did not take any approved vaccines. Whether the participants took the approved vaccines or not rests on the participants’ free-will. Although the number of participants in these subgroups was very small, we expected that the general trends without inference could be observed.

Although the data were not necessarily consistent among the assays (Ab titer, ntAb titer [pseudovirus], ntAb titer [cell-free]), there were general tendencies that participants who received EXG-5003 vaccines (both 5 µg and 25 µg groups) showed higher antibody levels than participants who did not receive EXG-5003 (placebo group; PBO) (Figure 6). The timing of blood sampling after the dosing of approved vaccines was not controlled, but there was no clear pattern of timing differences among the three groups, i.e., PBO, EXG-5003 (5 µg), EXG-5003 (25 µg).

**Figure 6.**
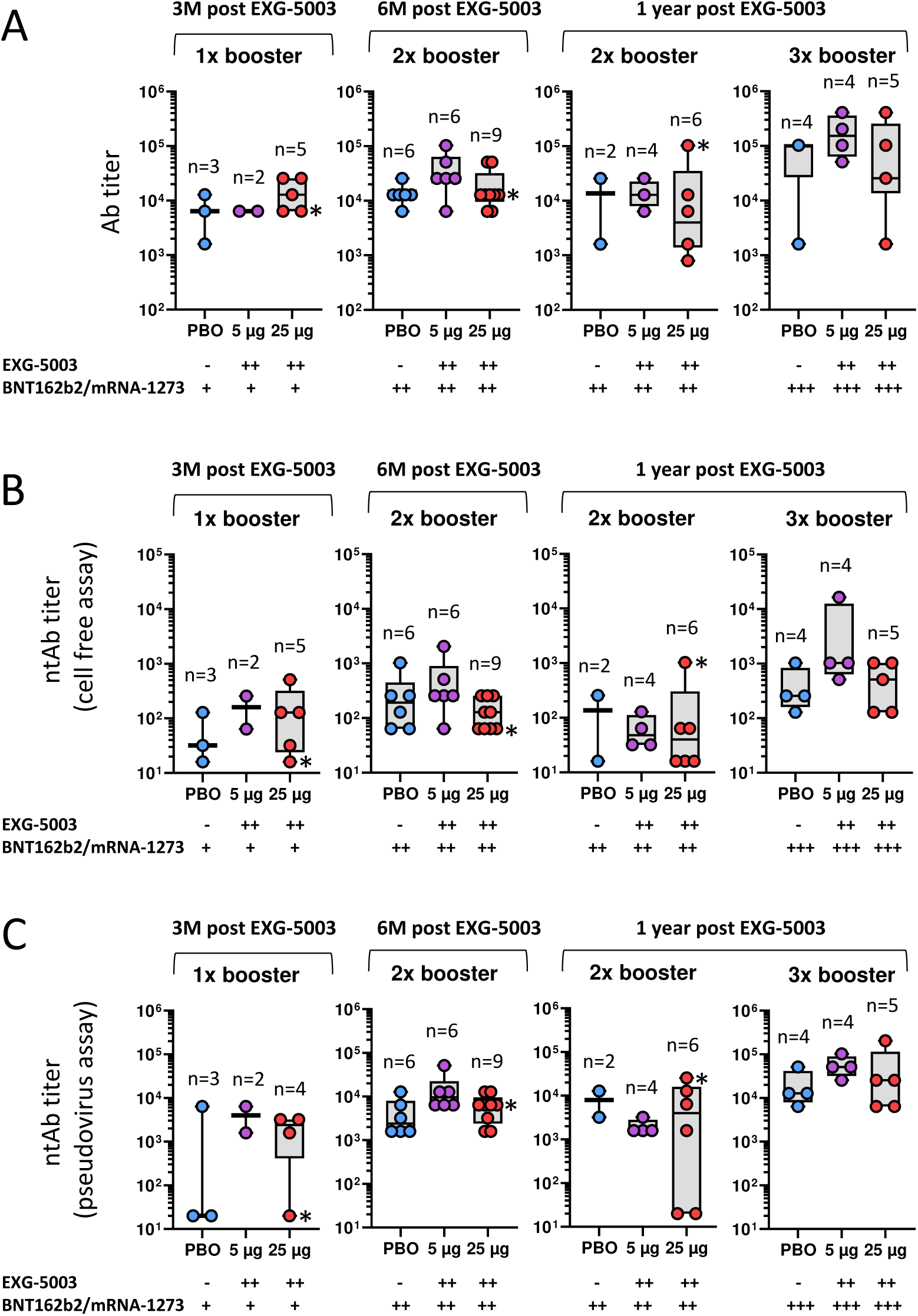
Long-term humoral immunity assessed by serum collected at 3 months, 6 months, and 1-year post-dosing with placebo (PBO), EXG-5003 (5 µg), or EXG-5003 (25 µg). Participants who received the approved mRNA vaccines (BNT162b2 or mRNA-1273) more than 10 days before the serum collection are shown. (A) Anti-RBD antibody titer. (B) Neutralization antibody titer by cell-free SARS-CoV-2 neutralization assay. (C) Neutralization antibody titer by the SARS-CoV-2 spike pseudo-typed lentivirus assay. * indicates individuals who became positive for antibodies against SARS-CoV-2 nucleoprotein, i.e., infected during 1-year follow-up period.

Notably, at 3 months post EXG-5003 vaccination when participants received only a single dose of BNT162b2 or mRNA-1273, participants who received prior EXG-5003 showed Ab titers higher than participants who did not receive EXG-5003 (PBO) (Figure 6A). Especially, EXG-5003 25 µg group showed the Ab titer comparable to participants who received two doses of BNT162b2 or mRNA-1273, but not received EXG-5003 (PBO) (Figure 6A). Similar trends were observed for ntAb titers assessed by both cell-free assay (Figure 6B) and pseudovirus assay (Figure 6C).

Taken together, EXG-5003 may have a priming effect for humoral immunity on subsequent heterologous vaccine administration.

### Long-term cellular immunity elicited by EXG-5003 following administration of licensed vaccines

The priming effect of EXG-5003 on subsequent heterologous vaccine administration was more clearly seen for cellular immunity. Figure 7A shows cellular immunity assayed by 15-mer peptide stimulation (i.e., CD4+, CD8+ T cells) at the 1-year follow-up (Table S5). Among participants who did not receive any approved vaccines by the 1-year follow-up, some individuals who received EXG-5003 vaccines possessed higher cellular immunity compared to the placebo group. These individuals were negative for SARS-CoV-2 nucleoprotein and, thus, were not infected throughout the study period. This trend was observed for participants who received two or three doses of an approved vaccine. The timing of blood sampling after the last vaccination (either EXG-5003 or approved vaccines) was around 10 months for no approved vaccine group, around 8 months for two doses of approved vaccines group, and around 1 month for three doses of the approved vaccines group (see the details in Figure 7 legend). The external controls were reproduced from Figure 5 for the comparison. For the convalescent group, blood sampling was done 20, 21, 21, and 22 days from the onset of COVID-19 symptoms. Blood sampling of BNT162b2 vaccine groups occurred before vaccination (BNT162b2 [pre]), 2 weeks after the first vaccination (BNT162b2 [1 dose]), and 2 weeks after the second vaccination (BNT162b2 [2 doses]) (Table S6). Two weeks after the vaccination is considered the time for peak cellular immunity ^11,12,25,26^.

**Figure 7.**
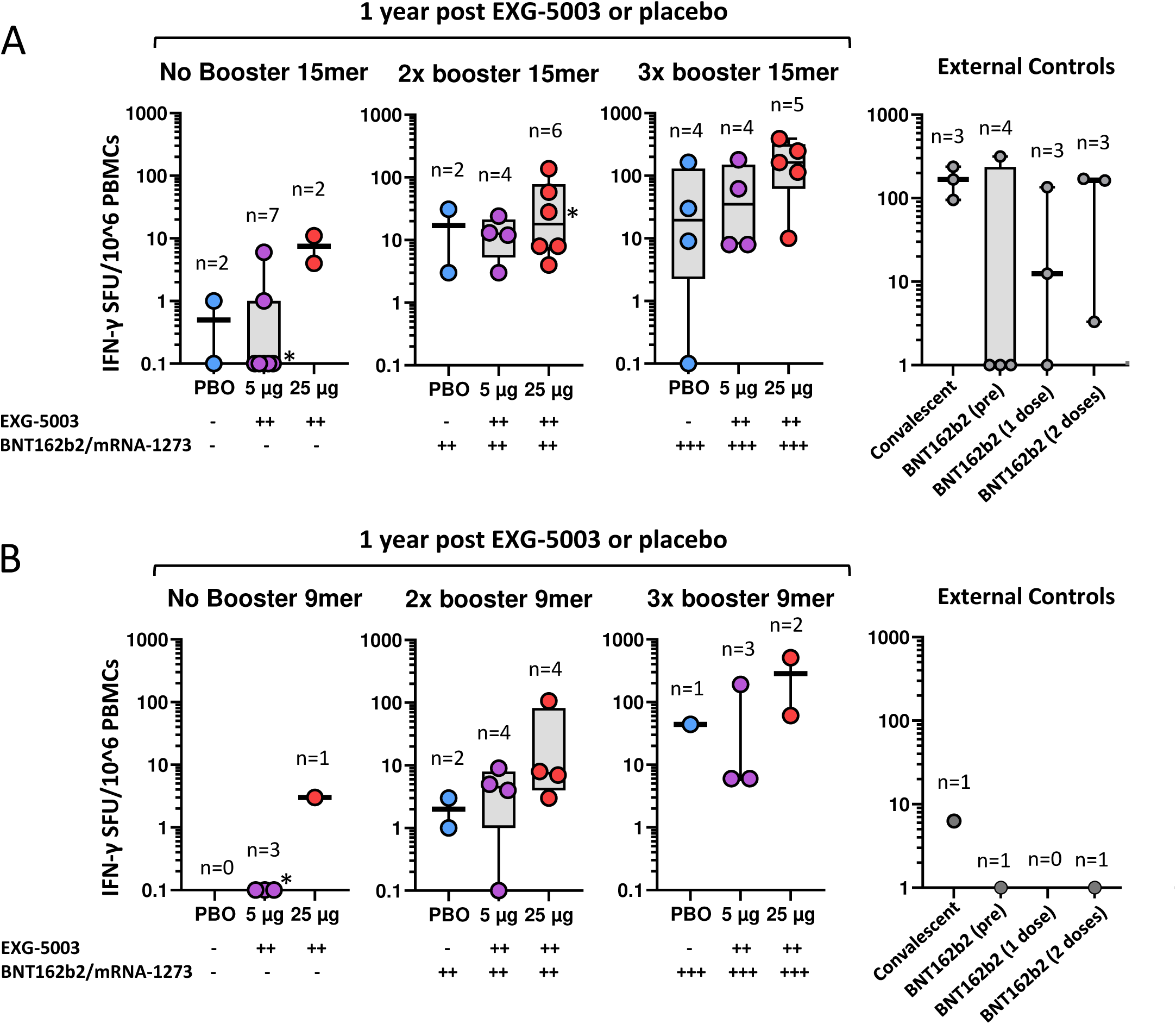
Long-term cellular immunity assessed by PBMCs collected at 1-year post-dosing with placebo (PBO), EXG-5003 (5 µg), or EXG-5003 (25 µg). (A) IFN-γ spot-forming units (SFU) per 10^6 PBMCs stimulated by 15-mer RBD peptide library. Participants were subdivided into three groups based on the timing of receiving the approved mRNA vaccines (BNT162b2 or mRNA-1273): No booster (days after receiving PBO [337 days], EXG-5003 5 µg [337 days], or EXG-5003 25 µg [337 days]), 2x booster (days after receiving the 2^nd^ dose of approved vaccines: PBO [205, 262 days], EXG-5003 5 µg [70, 190, 212, 253 days]; EXG-5003 25 µg [3, 205, 253, 270, 272, 283 days]), 3x booster (days after receiving the 3^rd^ dose of approved vaccines: PBO [46, 59, 63, 78 days]; EXG-5003 5 µg [13, 20, 21, 40 days]; EXG-5003 25 µg [20, 26, 61, 63, 68 days]). External controls (reproduced from Figure 5 for comparison): Convalescent (n=3, blood was collected 20, 21, 22 days after the onset of symptoms, respectively), BNT162b2 (pre) (n=4, before receiving 1st dose of BNT162b2), BNT162b2 (1 dose) (n=3, two weeks after receiving the 1st dose of BNT162b2), BNT162b2 (2 doses) (n=3, two weeks after receiving the 2nd dose of BNT162b2). * indicates individuals who became positive for antibodies against SARS-CoV-2 nucleoprotein, i.e., infected during 1-year follow-up period. (B) IFN-γ SFU per 10^6 PBMCs stimulated by 9-mer RBD peptide library. Participants were subdivided into three groups based on the timing of receiving the approved mRNA vaccines (BNT162b2 or mRNA-1273): No booster (days after receiving EXG-5003 5 µg [337 days], or EXG-5003 25 µg [337 days]), 2x booster (days after receiving the 2^nd^ dose of approved vaccines: PBO [205, 262 days], EXG-5003 5 µg [70, 190, 212, 253 days]; EXG-5003 25 µg [3, 205, 270 days]), 3x booster (days after receiving the 3^rd^ dose of approved vaccines: PBO [78 days]; EXG-5003 5 µg [13, 21, 40 days]; EXG-5003 25 µg [20, 25, 63 days]). External controls (for 9mer peptide library): Convalescent (n=1, blood was collected 20 days after the onset of symptoms, respectively), BNT162b2 (pre) (n=1, before receiving 1st dose of BNT162b2), BNT162b2 (2 doses) (n=1, two weeks after receiving the 2nd dose of BNT162b2). * indicates individuals who became positive for antibodies against SARS-CoV-2 nucleoprotein, i.e., infected during 1-year follow-up period.

Figure 7B shows cellular immunity assayed by 9-mer peptide stimulation (i.e., mainly CD8+ T cells) at the 1-year follow-up. There were fewer 9-mer than 15-mer peptide results as we prioritized the 15-mer peptide assays when blood samples were insufficient to run both. However, the priming effect of EXG-5003 was even more clearly seen compared to 15-mer peptide results (Figure 7A). Interestingly, both convalescent and twice BNT162b2 vaccinated individuals (BNT162b2 [2 doses]) in the control group showed very low 9-mer peptide results. This may be consistent with the notion that mRNA vaccine is a weak inducer of CD8+ T cells ^8–10^. The result shown in Figure 7B further highlights the potential of the EXG-5003 vaccine to enhance the CD8+ T cell response.

## Discussion

Synthetic mRNA vaccines have been enormously successful in combating the SARS-CoV-2 pandemic. However, it has also become clear that humoral immunity, and subsequently cellular immunity, rapidly wane, and thus, repeated vaccinations are recommended ^11,12^. Ideally, a vaccine that can provide long-term immune protection is desirable. Also, a vaccine that can provide not only humoral immunity, but strong cellular immunity, is desirable, because the cellular immunity is known to be more protective against variant forms of the virus. We attempted to address these issues with a novel intradermal c-srRNA vaccine platform, which is different from all currently licensed SARS-CoV-2 vaccines or those under development. For example, as far as we know, EXG-5003 is the only RNA vaccine that is administered intradermally and without LNP. Most likely due to this feature, the vaccine showed an excellent safety profile, including very few local and systemic adverse events.

As a standalone vaccine, the EXG-5003 vaccine did not induce Ab production. To some extent, this was expected from mouse studies^27^. On the other hand, mouse studies showed the induction of strong cellular immunity in both CD4+ and CD8+ T cells^27^, but the cellular immunity in humans was rather weak with a dose of up to 25 µg. However, it is worth noting that the level of cellular immunity achieved by EXG-5003 was comparable to traditional srRNA/saRNA vaccines, which are administered intramuscularly with LNP ^25,26^. Although the dose of traditional srRNA/saRNA vaccines was limited to 10 µg due to local and systemic adverse events, including grade 3 severity^26^, EXG-5003 can be dosed up to 25 µg without any severe adverse events. In fact, the clinical trial originally considered dose escalation of up to 100 µg, but this was not conducted due to insufficient induction of Ab production at 5 and 25 µg doses. Therefore, it remains to be seen whether higher doses of EXG-5003 can induce more robust cellular immunity as well as humoral immunity.

Surprisingly, EXG-5003 appears to provide long-term and enhanced priming effects on heterologous synthetic mRNA vaccines. For example, for T cell responses, assessed by FluoroSpot assays, in individuals who received EXG-5003 showed higher T cell immunity even around 10 months after receiving the licensed mRNA vaccines (i.e., booster) compared to individuals who did not receive EXG-5003. This priming effect was more pronounced in the FluoroSpot assays using a 9-mer peptide pool than those using a 15-mer peptide pool. Assuming that the former represents mainly CD8+ T cells and the latter represents CD4+ and CD8+ T cells^31^, the result is consistent with the preferential induction of MHC-I-mediated CD8+ T cells in mouse studies^27^. Furthermore, the priming effect of EXG-5003 was also consistent with the priming effect shown in mouse studies^27^. These results suggest that giving EXG-5003 as a primer vaccine provides immunological benefits to subsequent booster synthetic mRNA vaccines. As we showed in our mouse model, EXG-5003 induces the same immune enhancement effects even when used as a booster vaccine. Therefore, it is conceivable that a booster dose of EXG-5003 may confer immunologic benefits to people who have already received licensed mRNA vaccines. For such purposes, another vaccine, EXG-5008 — the same c-srRNA platform encoding for both RBD and nucleoproteins derived from SARS-CoV-2 and MERS-CoV ^27^, may be more suitable because it also has the benefit of more conserved nucleoproteins as antigens and intent to be a so-called pan-coronavirus vaccine.

One potential advantage of using the combination of c-srRNA vaccine and synthetic mRNA vaccine as heterologous prime-boost vaccines is that both are RNAs; thus, the advantages of RNA vaccines, such as rapid development and simple manufacturing, apply to both.

Furthermore, the c-srRNA vaccine does not use LNPs or nucleoside modifications, which are additional attractive features.

We acknowledge the limitations of our study. As a first-in-human trial of a novel c-srRNA vaccine platform, the number of participants was designed to be small. Furthermore, participants were allowed to take subsequent booster vaccines at their own will, resulting in even smaller numbers of participants in each subgroup of heterologous prime-boost vaccinations. Because of these small numbers, only the trends, without statistical inferences, were obtained.

In summary, we conducted a double-blind, placebo-controlled phase I/II trial to evaluate the safety, tolerability, and immunogenicity of EXG-5003, a two-dose, controllable self-replicating RNA vaccine against SARS-CoV-2. No safety concerns were observed with EXG-5003 administration. SARS-CoV-2 RBD antibody titers and neutralizing antibody titers were not elevated in either cohort, but elicitation of antigen-specific cellular immunity was observed in the EXG-5003 recipients in Cohort 2. Additionally, those who received an approved mRNA vaccine after receiving EXG-5003 showed higher cellular responses compared to equivalently vaccinated participants in the placebo group, suggesting a priming effect towards the cellular immunity of approved SARS-CoV-2 mRNA vaccines, and the potential applications of c-srRNA based vaccines against pathogens that depend on the cellular immunity protection.

## Materials and Methods

### EXG-5003 vaccine

The EXG-5003 vaccine was developed by Elixirgen Therapeutics, Inc. Its design and preclinical studies are described in the previous paper ^27^. In brief, this RNA vaccine platform is based on a self-replicating RNA (srRNA), also called self-amplifying RNA (saRNA or SAM), derived from the Venezuelan Equine Encephalitis virus. This RNA vaccine platform contains temperature-sensitive mutations, which makes it function at around 33°C (skin temperature) but not at around 37°C (core body temperature). Thus, it is called controllable self-replicating RNA (c-srRNA). The vaccine encodes the RBD of SARS-CoV-2 spike protein as an antigen. The EXG-5003 is intradermally delivered as naked RNA, i.e., without lipid nanoparticles. The EXG-5003 c-srRNA was diluted by Lactated Ringer’s solution at the clinical site and adjusted to the desired concentration.

### Trial design and participants

This was a single-center, phase 1/2, randomized, double-blinded, placebo-controlled trial conducted to evaluate the safety, tolerability, and immunogenicity of EXG-5003 in adults. The protocol, amendments, and overall oversight were approved by the Institutional Review Board for the Fujita Health University Hospitals (F-D-18) (jRCT2041210013, NCT04863131). The study was conducted in accordance with the Declaration of Helsinki.

The participants provided written informed consent before enrollment and randomization. Healthy adults aged 20 to 55 years who had a negative PCR test for SARS-CoV-2 and a negative anti-SARS-CoV-2 spike protein antibody were eligible. Key exclusion criteria included previous receipt of approved SARS-CoV-2 vaccine, symptoms consistent with COVID-19, uncontrolled cardiovascular, hematologic, respiratory, gastrointestinal, renal, and neuropsychiatric diseases, diabetes, autoimmune diseases, history of hepatitis B, hepatitis C, HIV infection, anaphylaxis or seizure, previous receipt of any SARS-CoV-2 vaccine, and ongoing pregnancy. Women of childbearing age were required to document a negative pregnancy test.

### Study procedures

The study enrolled two sequential cohorts, each consisting of 20 participants. Cohort 1 participants received 5 µg of EXG-5003 or placebo, whereas Cohort 2 participants received 25 µg of EXG-5003 or placebo. Participants were randomized in a 4:1 ratio to receive two 0.1 mL intradermal injections of either EXG-5003 or placebo (Lactated Ringer’s solution) to the posterior upper arm region, 28 days apart on Day 1 and Day 29 of randomization. Allocation of participants to the study arms were performed via a web-based interactive system by unblinded allocation staff, followed by preparation of EXG-5003 or placebo solution by unblinded pharmacists. The unblinded allocation staff and pharmacists who managed the study vaccine preparation had no role in participant assessment. The participants were followed for 52 weeks. The participants were allowed to receive SARS-CoV-2 vaccines that were approved in Japan (BNT162b2, mRNA-1273, ChAdOx1, NVX-CoV2373) after Day 57 during the follow-up period. In retrospect, most participants of this study received either BNT162b2 or mRNA-1273. For humoral immunity comparison, the following sera were used: one WHO standard serum; four convalescent individuals (serum was collected from COVID-19 patients hospitalized at Fujita Health University Hospital at 20, 21, 21, and 22 days after the onset of symptoms, respectively).

For cellular immunity comparison, blood samples were used from: three convalescent individuals (blood was collected from COVID-19 patients hospitalized at Fujita Health University Hospital at 20, 21, and 22 days after the onset of symptoms, respectively [patients are the same convalescent individuals described above]); four individuals (unrelated to this clinical trial) before receiving a 1st dose [BNT162b2 (pre)]; three individuals two weeks after receiving the BNT162b2 mRNA vaccine [BNT162b2 (1 dose)]; and three individuals two weeks after receiving the 2nd dose [BNT162b2 (2 doses)].

### Humoral immunity assays

#### Anti-RBD antibody

Antibody titers against SARS-CoV-2, including any antibodies that bind to the RBD, were measured by electrochemiluminescent (ECL) technology. The ECL is an immunoassay based on the excitation of a ruthenium sulfo-tagged antibody by an electrical current. Biotinylated RBD of the spike protein from SARS-CoV-2 (2019-nCoV) was captured onto an MSD Gold Streptavidin coated plate (ECL capable) for coating. This recombinant protein consisted of 234 amino acids corresponding to the RBD and had a predicted molecular mass of 28.7 kDa. A blocker solution was added to the wells and subsequently washed. The antibodies against the SARS-CoV-2 spike protein in the samples were captured onto the coated plate. After thorough washing of the wells to remove the unbound antibodies, sulfo-tagged anti-human IgG-FC was added to the wells to form a complex with the antibodies against the RBD. The excess unbound conjugate was then removed by further washing. The reaction was then developed by adding MSD read buffer, and the assay plate was read using an MSD ECL plate reader. The electrochemiluminescence signal generated was relative to the amount of antibodies against the RBD present in the samples tested.

#### Cell-free SARS-CoV-2 neutralization assay

Neutralizing antibody titers against the RBD of SARS-CoV-2 were measured by AlphaLISA technology. The AlphaLISA is an amplified luminescent proximity homogenous immunoassay based on the proximity of two types of beads: acceptor and donor beads. Briefly, all samples were initially diluted in assay diluent, diluted in MRD diluent, and transferred to an assay plate. They were then incubated with a mixture of acceptor beads (streptavidin-coated) and recombinant spike-RBD protein (biotinylated). During this step, the neutralizing antibodies bound to the spike-RBD protein. Lastly, a mixture of donor beads (anti-FLAG coated) and angiotensin-converting enzyme 2 (ACE2) protein (FLAG) were added to the reaction, by which non-neutralized spike-RBD protein would bind to the ACE2 receptor, bringing closer both acceptor and donor beads. Upon laser excitation, a photosensitizer present in the donor beads converted ambient oxygen to a more excited singlet state. The singlet oxygen channeled into the acceptor beads and reacted with a thioxene derivate in the acceptor beads, generating chemiluminescence at 370 nm that further activated a fluorophore (europium) embedded in the acceptor beads. The fluorophore subsequently emitted light at 615 nm, which was measured by the Envision reader. The amount of light generated in counts (cps) was inversely proportional to the amount of neutralizing antibodies against the RBD of the SARS-CoV-2 spike protein.

#### SARS-CoV-2 spike pseudotyped lentivirus assay

Neutralizing antibody titers against the RBD of SARS-CoV-2 were also measured by quasi-quantification of the antibody response in a titer approach. This cell-based assay using 293T-hsACE2 cells is based on the principle that binding between the SARS-CoV-2 spike protein and the cellular receptor ACE2 protein is necessary for cellular infection. In the presence of neutralizing antibodies, the spike protein function is affected, and therefore, the binding between the spike and ACE2 proteins is negatively affected. This assay uses luciferase reporter virus particles (RVPs).

Samples were initially diluted in cell culture media and transferred to a 96-well plate. A known quantity of RVPs (or pseudovirus) was added and incubated to allow antibody binding. A set quantity of 293T-ACE2 cells was added, and plates were incubated for 48 hours. After the incubation period, a luciferase substrate was added to each well, and relative light unit (RLU) values were read using luminescence settings. The amount of RLUs was inversely proportional to the amount of neutralizing antibodies against the SARS-CoV-2 spike protein.

The inhibitory rates were calculated based on RLU values and used for assessing antibody and serum neutralization activities. The endpoint titers were calculated by serial dilutions of the samples.

### Cellular immunity assays

#### FluoroSpot assay

This FluoroSpot (FS) assay was conducted to detect and quantify interferon gamma (IFN-γ) and interleukin 4 (IL-4) secretion by antigen-stimulated human PBMCs. PBMCs were isolated from three groups, including participants of this trial, convalescent patients who were hospitalized and recovering from COVID-19, and participants of a separate study that enrolled individuals receiving the BNT162b2 mRNA vaccine. SARS-CoV-2 peptide pools (9-mers and 15-mers) and controls (Phytohemagglutinin, Dimethyl sulfoxide, and CPI [protein antigens of CMV, Parainfluenza and Influenza viruses]) were used as stimulants.

Peptide pools used to stimulate PBMC are described in a previous paper ^27^. In brief, the 15-mer peptide pool is a pool of 53 peptides derived from a peptide scan (15-mers with 11 amino acid overlap) through the RBD of SARS-CoV-2 (an original Wuhan strain) [JPT Peptides: PepMix SARS-CoV-2 (S-RBD) PM-WCPV-S-RBD-2]. The 9-mer peptide pool is a pool of 215 peptides derived from a peptide scan (9-mers with 8 amino acid overlaps) through the RBD of SARS-CoV-2 (an original Wuhan strain). The 9-mer peptide pool was custom-made by JPT Peptides. It is known that the 15-mer peptide pool induces both CD8+ T cell response and CD4+ T cell response reliably, whereas the 9-mer peptide pool induces primarily CD8+ T cell response; however, the CD4+ T cell response is also induced less reliably^31^.

FluoroSpot assay plates were washed with PBS and blocked with CTL media for ≥ 30 mins at 37°C. Post-thaw PBMCs were seeded at 4×10^5 cells per well (100 µL) and stimulated in triplicate with one of the following five different stimulants: 1 µg/mL of 9-mer SARS-CoV-2 or 15-mer SARS-CoV-2 peptide pools, 5 µg/mL CPI, 3 µg/mL PHA, or DMSO (negative control). The final combined volume of cells and stimulant was 200 µL per well. Following 48 hours of incubation, plates were washed and incubated with Fluorescein isothiocyanate (FITC)-labeled anti-IFN-γ or biotinylated anti-IL-4 detection antibodies for 2 hours at room temperature in the dark. Plates were then washed and incubated for 1 hour at room temperature in the dark with either anti-FITC Alexa Fluor 488 tertiary antibody for detecting IFN-γ or streptavidin CTL Red for detecting IL-4. Plates were finally washed with water and placed in the dark to dry overnight. Within 24-36 hours after development, plates were scanned using the CTL. The numbers of spot-forming cells (SFCs) were normalized to spots per million cells to yield spot-forming units (SFUs).

#### Safety assessment

Local and systemic adverse events, as defined in the FDA guidance for preventive vaccine clinical trials, were solicited at each follow-up visit (Days 1, 8, 15, 29, 36, 43, 57, 92, 183, 365). Laboratory values including complete blood count, basic metabolic panel, liver function tests, and urinalysis were assessed on select follow-up visits (Days 1, 8, 29, 36, 57).

#### Statistical analysis

Data were analyzed using R (version 4.1.0) and GraphPad Prism (version 10.0.2). Sample size was determined in advance primarily for feasibility considerations rather than through statistical inferential testing. Due to a small sample size, only descriptive statistics were used and presented in this paper.

## Supplementary materials

The following supporting information can be downloaded at: www.mdpi.com/xxx/s1, Table S1: Antibody titers; Table S2: Antibody titers of external controls; Table S3: Neutralizing Antibody titers (Cell-free assays); Table S4: Neutralizing Antibody titers (Pseudovirus assays); Table S5: Fluorospot assay results at 1-year post-EXG-5003 vaccination; Table S6: Fluorospot assay results for external controls.

## Author contributions

TK, MT, HF, MaK, YD conducted the clinical trial at Fujita Health University. MiK coordinated regulatory activities. MSHK, TA, MA, MB, HY, PR, EL, JK, HY, ACK, NPW, AA were involved in EXG-5003 vaccine development. HY conducted data management. IT, ST, AS conducted planned statistical analyses. MSHK and YD conducted post-hoc statistical analyses and drafted the manuscript.

## Institutional review board statement

The protocol, amendments, and overall oversight were approved by the Central Institutional Review Board for the Fujita Health University Hospitals (F-D-18) (jRCT2041210013, NCT04863131). The study was conducted in accordance with the Declaration of Helsinki.

## Informed consent statement

Informed consent was obtained from all participants involved in the study.

## Data availability statement

The data presented in this study can be found in Figures, Tables, and Supplemental Tables.

## Acknowledgments

We thank Allie Amick, Ph.D. for editing the manuscript.

